# Cardiac imaging of aortic valve area from 26,142 UK Biobank participants reveal novel genetic associations and shared genetic comorbidity with multiple disease phenotypes

**DOI:** 10.1101/2020.04.09.20060012

**Authors:** Aldo Córdova-Palomera, Catherine Tcheandjieu, Jason Fries, Paroma Varma, Vincent S. Chen, Madalina Fiterau, Ke Xiao, Heliodoro Tejeda, Bernard Keavney, Heather J. Cordell, Yosuke Tanigawa, Guhan Venkataraman, Manuel Rivas, Christopher Ré, Euan Ashley, James R. Priest

**Affiliations:** Department of Pediatrics, Division of Pediatric Cardiology Stanford University School of Medicine, Stanford, 94304, CA; Department of Computer Science, Stanford University, Stanford, CA; Center for Biomedical Informatics Research, Stanford University, Stanford, CA; Department of Electrical Engineering, Stanford University, Stanford, CA; Division of Cardiovascular Sciences, Faculty of Biology, Medicine and Health, The University of Manchester, Manchester, UK; Division of Medicine, Manchester University NHS Foundation Trust, Manchester Academic Health Science Centre, Manchester, UK; Population Health Sciences Institute, Faculty of Medical Sciences, Newcastle University, Newcastle upon Tyne, UK; Department of Biomedical Data Science, Stanford University, Stanford, CA; Department of Medicine, Stanford University, Stanford, CA

**Keywords:** aortic valve area, GWAS, polygenic score, PheWAS, genetic comorbidity

## Abstract

The aortic valve is an important determinant of cardiovascular physiology and anatomic location of common human diseases. From a sample of 26,142 European-ancestry participants, we estimated functional aortic valve area by planimetry from prospectively obtained cardiac MRI sequences of the aortic valve. A genome-wide association study of aortic valve area in these UK Biobank participants showed two significant associations indexed by rs71190365 (chr13:50764607, *DLEU1, p*=1.8×10^−9^) and rs35991305 (chr12:94191968, *CRADD, p*=3.4×10^−8^). From the GWAS findings we constructed a polygenic risk score for aortic valve area, which in a separate cohort of 311,728 individuals without imaging demonstrated that smaller aortic valve area is predictive of increased risk for aortic valve disease (Odds Ratio 0.88, *p*=2.3×10^−6^). After excluding subjects with a medical diagnosis of aortic valve stenosis (remaining n=310,546 individuals), phenome-wide association of >10,000 traits showed multiple links between the polygenic score for aortic valve disease and key health-related comorbidities involving the cardiovascular system and autoimmune disease. Genetic correlation analysis supports a shared genetic etiology with between aortic valve size and birthweight along with other cardiovascular conditions. These results illustrate the use of automated phenotyping of cardiac imaging data from the general population to investigate the genetic etiology of aortic valve disease, perform clinical prediction, and uncover new clinical and genetic correlates of cardiac anatomy.

## INTRODUCTION

The geometry of the aortic valve underlies cardiac structure and function relationships, including and fluid dynamics, response to stress and valvular pathology ^1^. Aortic valve stenosis (AS) is a common cardiovascular condition characterized by narrowing of the functional orifice of the aortic valve with an ensuing obstruction to ejection of blood from the left ventricle and increased wall stress. In adult cohorts AS is the most frequent valvular disease ^2^, with prevalence estimates around 0.3-0.5% in the general population, and markedly higher estimates in older individuals (e.g., circa 7% in subjects >65-year old subjects) ^3^. As the natural history of adult-onset AS involves a relatively asymptomatic course followed by declining health after symptoms appear ^4-7^, there is a strong therapeutic potential for early detection of disease^8^.

A complex heritability pattern for AS involving multiple genes has been suggested ^9^, and larger genome-wide association studies (GWAS) have linked novel genomic loci to disease risk ^10,11^. However additional molecular genetic mechanisms of AS remain to be discovered. The advent of large population-based datasets that combine genetic data with cardiac imaging data such as the UK Biobank ^12^ offers unprecedent opportunities to investigate heritable factors underlying AS pathogenesis to ultimately propose tools for the early identification of at-risk individuals.

While diagnosis of aortic valve stenosis may be derived from a combination of findings from physical exam, auscultation, echocardiography or functional data obtained during cardiac catheterization or surgery, recent findings support the clinical utility of software-based methods for magnetic resonance imaging (MRI) analysis ^13^. Novel magnetic resonance imaging (MRI)-based techniques to automatically distinguish between bicuspid and normal (tricuspid) aortic valve have potential to translate into clinical applications ^14,15^, and to facilitate biomedical research through the use of large biobanks^16^. Here, we use planimetric measures of the aortic valve functional area obtained through an automated approach for cardiac MRI data ^16^ as a proxy for valvular function, to conduct a genome-wide association study on 26,142 individuals of European ancestry from the UK Biobank. We compute polygenic scores and perform a phenome-wide association study to search for potential links between the genetic basis of aortic valve size and other health-related phenotypes.

## RESULTS

### Participants and aortic valve measurements

MRI-derived planimetric measurements of the aortic valve area (Figure 1) were obtained for 26,142 European-ancestry subjects (51.5% females, mean (SD) age: 55.06 (7.4) years, mean (SD) aortic valve size: 435.8 (134.5) mm2) with genotyping data available through the UK Biobank. As displayed on Figure 1, the estimated aortic valve area indexed to body surface area was calculated at 2.28 cm^2^/m^2^, concordant with aortic valve area measured directly at autopsy in a population of 4,803 adults ^22^.

**Figure 1.**
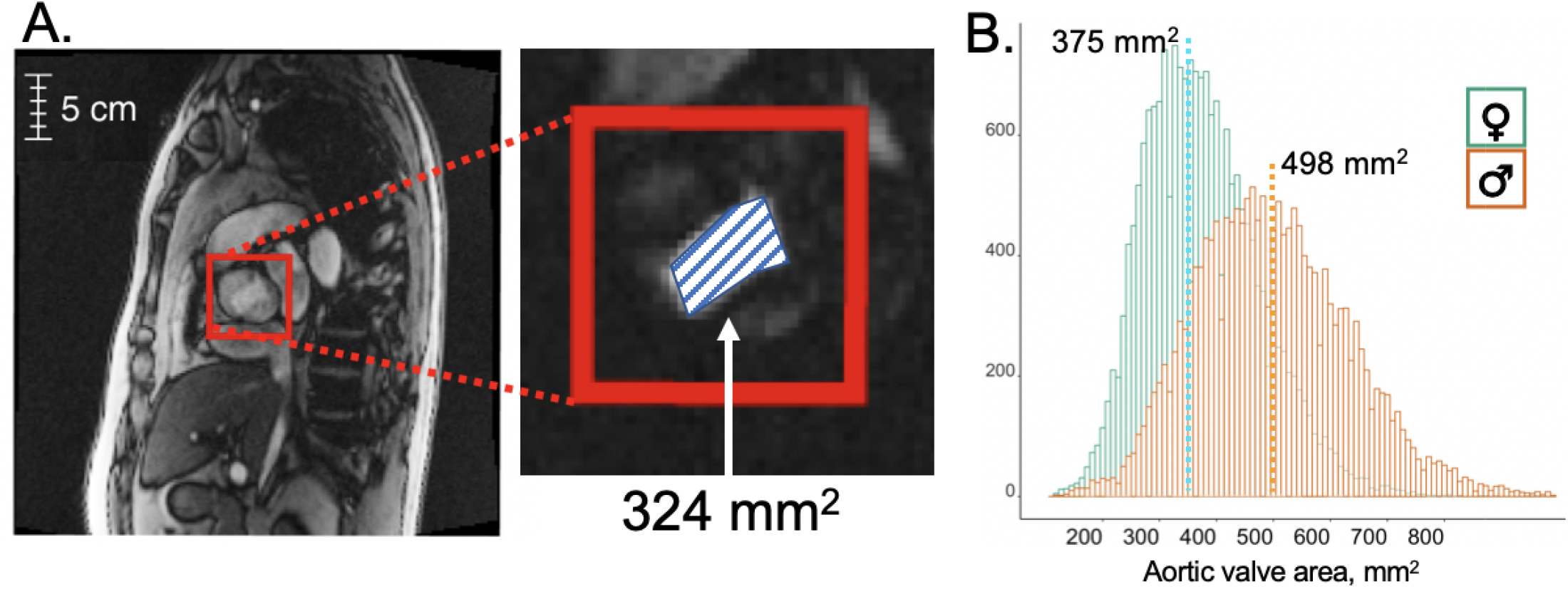
Aortic valve size measurement from MRI sequence data. A: MRI frames for CINE and MAG series in an oblique coronal view of the thorax centered upon an *en face* view of the aortic valve at sinotubular junction (red boxes). B: Distribution of aortic valve size measurements in males and females. Estimated aortic valve area indexed to body surface area is calculated at 2.28 cm^2^/m^2^ which is concordant with aortic valve area measured directly at autopsy in a population of 4,803 adults ^22^.

### GWAS of aortic valve size

Aortic valve measurements were submitted to genome wide association testing (GWAS) for over 4.1 million markers using standard linear regression. A GWAS of aortic valve size within the MRI cohort (n=26,142) revealed two genome-wide significant associations (Figure 2); summary statistics did not show evidence of inflation (genomic inflation factor (lambda): 1.02). The two observed associations were an intronic variant at the Deleted In Lymphocytic Leukemia 1 gene (*DLEU1*) (chr13:50753830-50792591, lead variant: rs71190365 (chr13:50764607_C_CT), minor allele frequency (MAF)=30.5%, beta (B)=6.91, standard error (SE)=1.15, *p*=1.8×10^−9^), and a variant located on an intron of the CASP2 And RIPK1 Domain Containing Adaptor With Death Domain gene (*CRADD*) (chr12:94184082-94201279, lead variant: rs35991305 (chr12:94191968_T_TG), MAF=40%, B=-5.87, SE=1.06, *p*=3.4×10^−8^) (Figure 3). In addition, a region on chromosome 17 showed a marginally significant signal led by a marker on the Golgi SNAP Receptor Complex Member 2 (*GOSR2*) (chr17:44967530-45244074, lead variant: rs17608766 (chr17:45013271_T_C), MAF=15%, B=7.83, SE=1.44, *p*=5.6×10^−8^).

**Figure 2.**
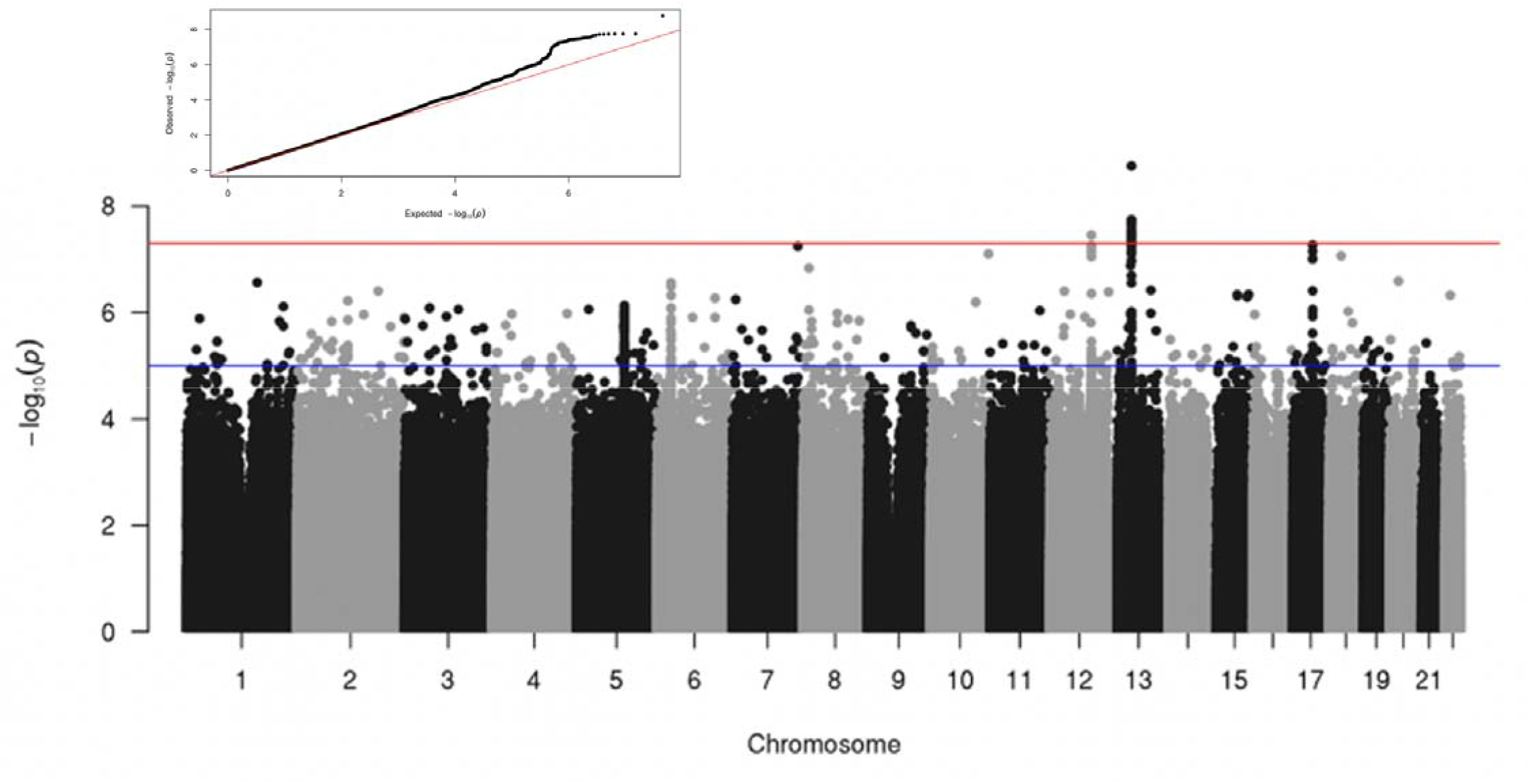
Manhattan and QQ-plot from the GWAS of MRI-derived aortic valve area (n=26,142). A quantile-quantile (QQ) plot of the p-value distribution is shown on the top left corner. The red horizontal line indicates the conventional multiple testing correction for GWAS (*p*=5×10^−8^), and the blue line shows suggestive associations (*p*=5×10^−5^).

**Figure 3.**
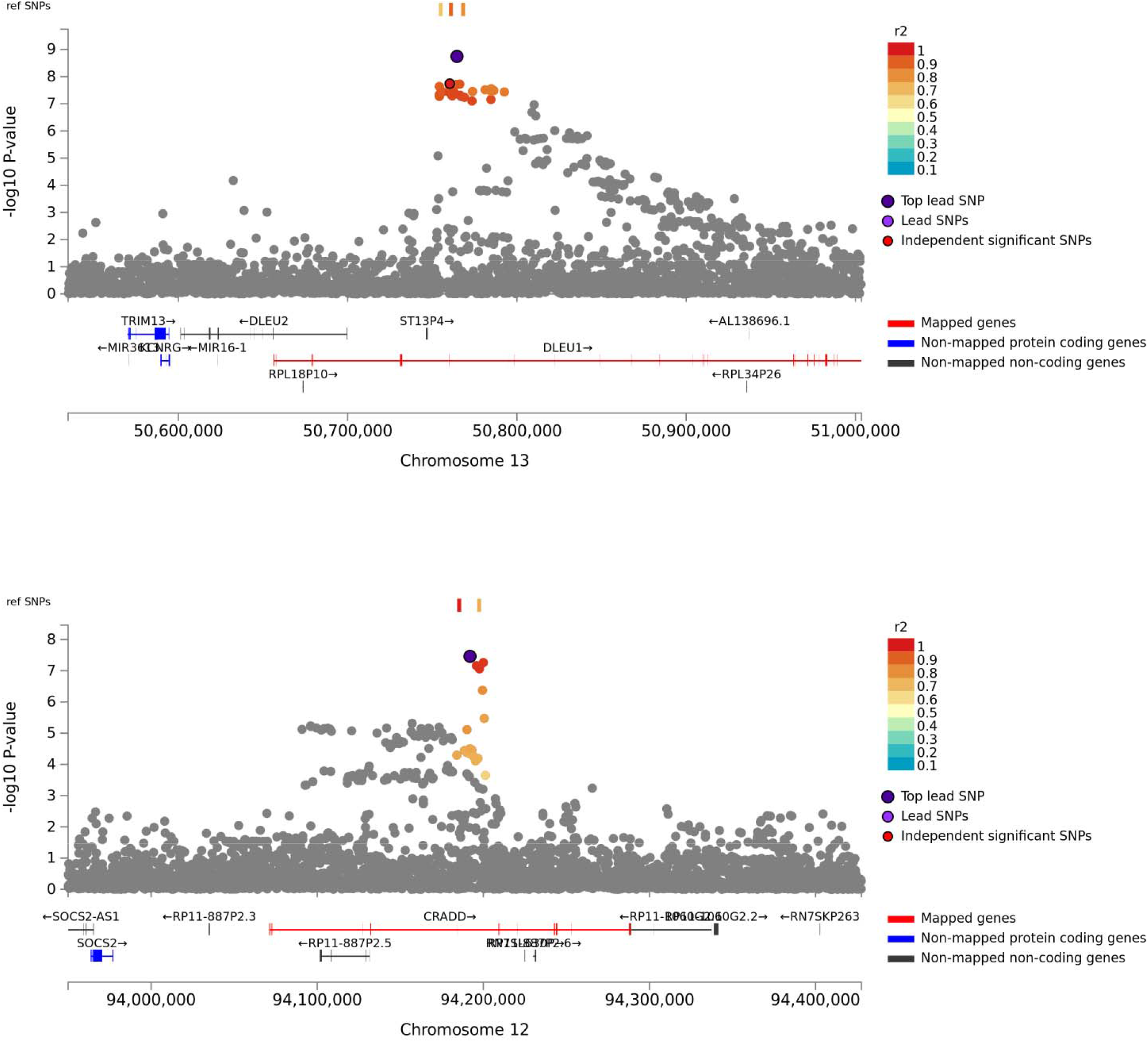
Locus Zoom plots for two genome-wide significant loci identified through the aortic valve area GWAS plots showing recombination rates and overlapping phenotype associations from the GWAS literature are included on Supplementary Figure S5.

Additional insights on putative candidate genes and functional annotation and mapping were obtained through the FUMA platform^17^. First, set-based analysis of 19,148 protein-coding genes on the genome-wide aortic valve area results performed through MAGMA^18^ showed seven genes significant at *p*=0.05/19148=2.611e-6, in the following order: *CRADD* (chr12:94071151-94288616, Z=6.09, p=5.6e-10), *DLEU1* (chr13:50656307-51297372, Z=5.49, p=2e-8), *BRAP* (chr12:112079950-112123790, Z=4.85, p=6.02e-7), *IQCH1* (chr15:67547138-67794598, Z=4.7628, p=9.55e-7), *GOSR2* (chr17:45000483-45105003, Z=4.7099, p=1.2394e-06), *RP11-162P23*.*2* (chr12:112191694-112229222, Z=4.6684, p=1.5175e-06) and *FER* (chr5:108083523-108532542, Z=4.6156, p=1.96e-6) (Supplementary Figure S1).

In addition, data from expression quantitative trait loci (eQTL) suggested that the genome-wide significant locus on chromosome 12 may impact the expression of *MRPL42* and *SOCS2*, whereas the significant region on chromosome 13 might involve six genes (*DLEU1, DLEU7, EBPL, KCNRG, RCBTB1* and *TRIM13*).

### Aortic valve size polygenic score and PheWAS

From the summary statistics of the aortic valve area GWAS, we then created a polygenic score (PGS) for aortic valve area for all European-ancestry subjects as the weighted sum of alleles associated with aortic valve area (see Methods). Amongst the UK Biobank participants of European ancestry who did not have MRI data (n=310,546) we identified 732 participants with aortic valve disease as defined by inpatient diagnosis codes ^19^. For each standard deviation decrease in the PGS for aortic valve area there was a strong and statistically increased risk for clinically defined aortic valve disease (beta=0.13, SE=0.03, *p*=2.3×10^−6^).

To better understand the clinical correlates and predictors of aortic valve area, we conducted a phenome-wide association study (PheWAS) relating the genome-wide PGS with 2,976 phecodes ^20^ and phenotypes from the Global Biobank Engine (GBE) ^21^, applied to the subset of healthy controls without MRI images (n=310,546 individuals). As displayed on Figure 4, a higher PGS for aortic valve size was related to a lower risk for multiple types of diseases: digestive (e.g., celiac disease), metabolic and endocrine (e.g, diabetes, hypothyroidism), circulatory (e.g., angina pectoris, ischemic heart disease) and respiratory (e.g., bronchitis). It also showed the inverse association (risk-increasing) for foot deformities and disorders of iron metabolism. PheWAS results for the two lead markers on chromosomes 12 and 13 are included on Supplementary Figure S2, Supplementary Table S1 and Supplementary Table S2.

**Figure 4.**
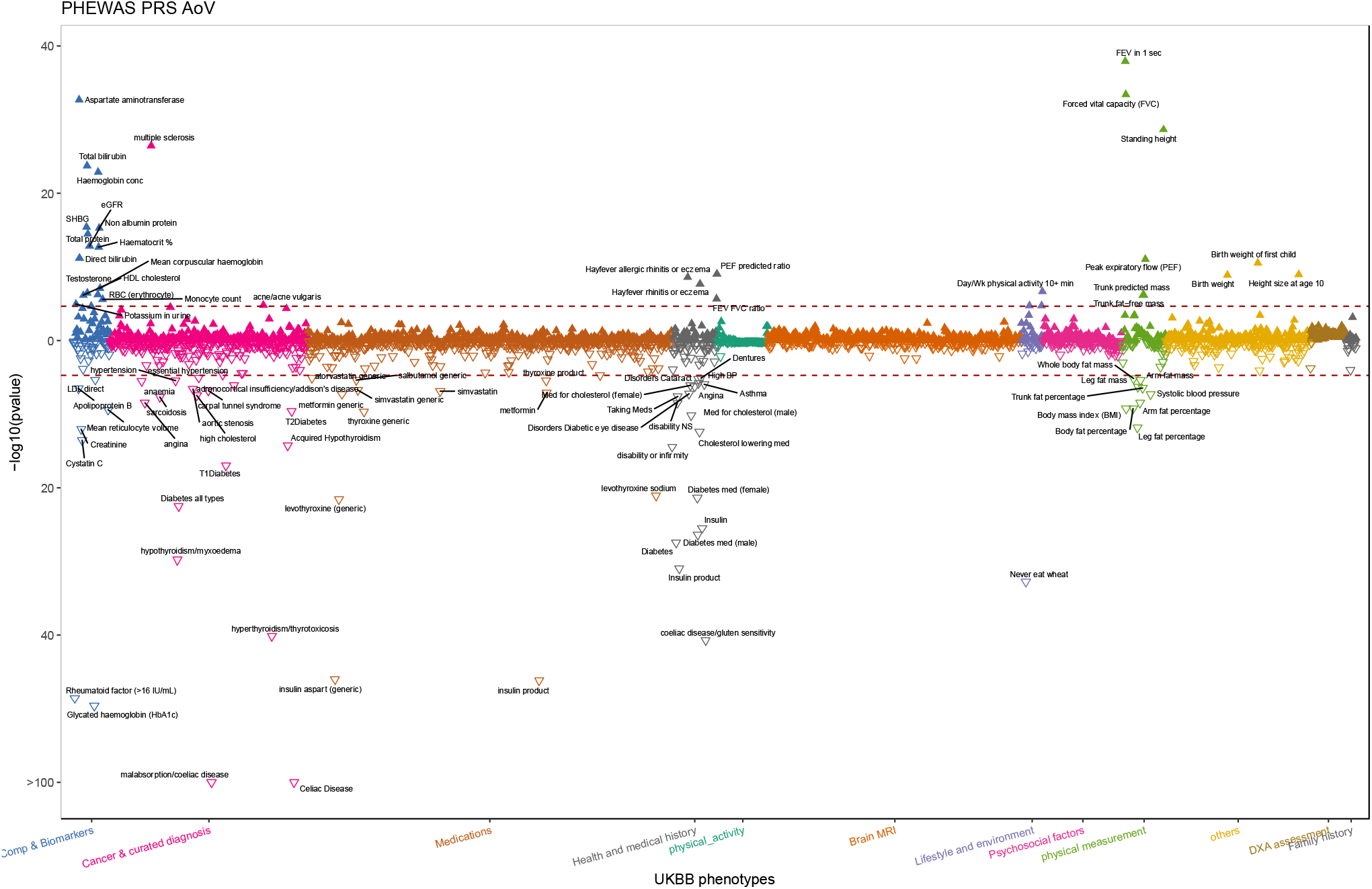
Phenome-wide association results for the aortic valve size polygenic score using ICD-10 codes mapped to phecodes. Additional results from the Global Biobank Engine ^21^ are included in Supplementary Figure S4. Phenotypes are colored by category (shown on the horizontal axis) and each triangle points up or down depending on the direction of effects (positive or negative associations).

Finally, we performed genome-wide correlation analysis of aortic valve area with 597 UK Biobank GWAS results along with 258 independent studies using the LDHub tool ^23,24^. Supplementary Figure 2 shows significant correlations at a false discovery rate of *p*=0.05 separately for published research studies (non-UK Biobank) and UK Biobank GWAS results (http://www.nealelab.is/uk-biobank, which is complicated by phenotypic and sample overlap between the lab GWAS traits and e.g., aortic valve size comorbidities in subjects from the current MRI study). After correction for multiple testing, the correlation with UK Biobank summary statistics indicated that alleles associated with larger aortic valve size show a positive correlation with birthweight and a number of functional measures including pulmonary function (forced vital capacity, forced exploratory volume in 1-second), hand grip strength, and walking pace similar to the results from the PheWAS. The same alleles displayed a negative correlation (apparent protective effect) with a personal and family history of diabetes and heart disease, visual problems, and anthropometric measures linked to cardiometabolic disease (fat percentage, waist circumference and body mass index). Amongst the independent 258 genetic correlation coefficients, alleles related to larger aortic valve size overlap with the genetic determinants of birthweight ^25,26^ (Supplementary Table S3).

## DISCUSSION

In this work, computationally derived MRI measures of aortic valve area were screened to find genetic associations amongst European-ancestry subjects from the UK Biobank. Through GWAS of common variants, two loci were identified: a signal indexed by an intronic *DLEU1* variant (rs71190365, chr13:50764607, *p*=1.8×10^−9^) and a second region led by a marker intronic within the *CRADD* gene (rs35991305, chr12:94191968, *p*=3.4×10^−8^). A lower polygenic score for aortic valve area was predictive of clinically-defined aortic valve disease in the set of UK Biobank subjects without an MRI scan available (2,000 and 310,546 subjects with and without a history of aortic valve disease, respectively). Finally, in individuals without a diagnosis of aortic valve disease (n=310,546), PheWAS results suggested links between genetic markers related to aortic valve size and cardiovascular complications.

Supporting our computational approach to measurement of aortic valve area, the two novel loci identified through GWAS findings have been described previously in the literature. The identified region on chromosome 12 displays an intronic variant in the *CRADD* gene as a top marker. Variation mapping to *CRADD* has been previously related to coronary artery calcification ^27^, a phenotype strongly linked to calcific aortic valve disease and aortic stenosis ^28^. The PheWAS data for rs71190365 indicate a link to known cardiac comorbidities such as forced expiratory volume and body mass, which may represent a pleiotropic effect—decreased area of the aortic valve is a determinant of cardiac output which governs cardiac output and exercise capacity ^29^. Functional eQTL information indicates that this GWAS signal may involve *SOCS2* as causal gene, which has been implicated in early body growth processes ^30,31^. In this regard, PheWAS results on Supplementary Figure S2 also suggest a role for the lead SNP in body mass and height. The second locus retrieved from the GWAS is indexed by an intronic SNP on *DLEU1* which has been linked to echocardiographic measures of aortic root diameter ^32^.

Importantly, the polygenic score derived from genome-wide alleles related to aortic valve area was associated with a clinical history of aortic valve disease, suggesting that the genetic signals from population-based cardiac MRI measurements may predict risk for health and disease. Exploring this score via a PheWAS, the same PGS estimates were linked to higher risk for several conditions (notably, multiple sclerosis) and lower risk for celiac disease, angina and cardiovascular conditions. Aortic valve dysfunction is a common feature of Turner syndrome along with autoimmune celiac disease ^33,34^, while aortic valve disease may also accompany component autoimmune phenotypes of Multiple Sclerosis ^35,36^. However causal relationships with aortic valve area are not revealed by these analyses, the observed associations between a PGS and a second phenotype may represent some aspect of shared genetic architecture between two traits or clinically correlated epiphenomena. Validation of this PGS in other large cohorts is needed to confirm the predictive characteristics for aortic valve disease and other clinically relevant phenotypes identified here.

The findings from the PheWAS are supported by the genetic correlation analyses from the UK Biobank studies; although not passing a strict multiple testing correction through false discovery rate, they are internally consistent with the 258 independent-trait analyses. Aortic valve size alleles also display a negative enrichment (apparent protective effect) for coronary artery disease (rg=-0.211, z=-3.0126, p=0.0026 ^37^) and type 2 diabetes (rg=-0.2347, z=-2.2619, p=0.0237 ^38^), and a positive correlation with forced vital capacity (rg=0.1972, z=2.5913, p=0.0096 ^39^, and rg=0.2481, z=2.2151, p=0.0268 ^40^). These findings are also likely to represent pleiotropy—lung function and aortic valve size are tightly linked to body size, while aortic valve flow is a functional determinant of blood flow in the coronary arteries ^41^.

Some limitations of this study deserve mention. First, since the UK Biobank is not a hospital-based sample, the range of aortic valve sizes may be narrower than what is observed in clinical datasets, and it is certain that selection and survivorship biases have influenced the GWAS results ^42^. While the aortic valve measurements were almost entirely automated, systematic errors related to the underlying computational approach may introduce inaccuracies. However, the fact that the findings are biologically relevant likely reflects that true genetic effects were present in the observed signals.

Overall, the outcomes suggest novel candidate loci as determinants of aortic valve size in the general population and indicate a shared genetic architecture with different traits across the health-disease spectrum. Leveraging imaging and diagnostic information from large-scale public records offer an unprecedented opportunity to study the genetic architecture of variability in cardiac morphology and its link to aortic valve disease.

## METHODS

### Ethics statement

De-identified data from the UK Biobank was used. Ethical approval for the UK Biobank was granted by the NHS National Research Ethics Service (ref: 11/NW/0382).

### Participants and measures

Genetic and phenotypic information was extracted from the UK Biobank cohort^12,43^, a detailed prospective study of >500,000 individuals aged 40-70 at enrollment. Recruitment was conducted between 2006 and 2010; the study collected extensive phenotypic information about the participants, including medical histories ascertained by healthcare professionals through verbal interview, electronic health records (EHRs) with inpatient diagnosis codes from the International Classification of Disease 9 and 10 (ICD-9 and 10) and Office of Population and Censuses Surveys (OPCS-4) diagnosis codes. Discovery analyses included the largest UK Biobank subset of unrelated individuals of European-ancestry with both MRI and genetic data available at the time of manuscript preparation (up to 26,169 participants in total; 51% female; mean age of 55.1 years). Follow-up polygenic score analyses included unrelated, European-ancestry individuals who were not included on the previous stage (732 cases and 310,546 controls, % female, mean age), and phenome-wide association tests were conducted after excluding the aortic valve disease cases from the polygenic score stage (310,546 subjects without a history of aorthopaty).

### Estimation of aortic valve functional area

Aortic valve functional area was derived from a planimetric estimate of the aortic valve orifice from a set of MRI sequences targeted to the aortic valve ^44^. In common clinical practice, the aortic valve functional area may be computed using the Gorlin continuity equation which incorporates Doppler echocardiographic measurements obtained proximal and distal to the aortic valve with the principle of conservation of energy ^45^. In clinical studies, planimetric measurement of aortic valve area from cardiac MRI is strongly concordant with echocardiographic measures derived from the Gorlin continuity equation ^46,47^. For each subject with available cardiac imaging from the first UK Biobank data, three sequences of phase-contrast MRI of the aortic valve with *en face* views at the sinotubular junction were obtained: magnitude (MAG), raw anatomical (CINE) and velocity encoded (VENC). Images were preprocessed to localize the aortic valve to a 32×32 grid and align all frames by peak blood flow in the cardiac cycle (Figure 1). A more detailed description of the initial protocol, including supporting videos, is presented by Fries et al ^16^.

### Genotyping data and association tests

The UK Biobank data release available at the time of analysis included genotypes for 488,377 participants, obtained through either the custom UK Biobank Axiom array or the Affymetrix Axiom Array. Genotypes imputed to the Haplotype Reference Consortium ^48^ and the combined UK10K/1000 Genomes panels were retrieved from the UK Biobank data showcase. Only variants with a minor allele count (MAC) greater or equal to 20 in the entire MRI dataset and an empirical-theoretical variance ratio (MaCH’s *r*^2^) threshold above 0.3 were included. Family relatives and non-European ancestry participants were excluded based on genetic features provided with the latest UK Biobank data release ^12^, and a subset totaling 26,169 individuals with cardiac MRI was included in the genetic association tests described next. Association tests were performed using linear regression PLINK2 ^49^, including gender and adjusted body surface area as covariates.

### Phenome-wide association study

Phenome-wide association studies (PheWAS) were conducted to identify associations between relevant genotypes and up to 2,976 clinical diagnoses, represented as phecodes ^20^, through the Global Biobank Engine (GBE) ^21^. The GBE is a tool that enables exploration of the relationship between genotype and phenotype in cohorts such as the UK Biobank, and supports browsing for results from GWAS, PheWAS and other genetic association tests. Additionally, when relevant, PheWAS of up to 7,569 traits curated through PHESANT ^50^ on the UK Biobank was also performed. PHESANT is a dedicated software tool for large-scale UK Biobank phenome screenings implemented as an R package, which provides an automated processing workflow to determine variable coding and standardization of continuous, integer and both single- and multi-value categorical fields. Variables standardized through PHESANT on the largest subset of unrelated European-ancestry subjects not included in the MRI analysis (up to 310,546 subjects) were then submitted to PheWAS using a custom R script, with linear regression for continuous and integer variables, and logistic regression for re-coded categorical variables. For all PheWAS reported here, phenotypes with less than 100 individuals (for continuous traits) or less than 100 cases and 100 controls (for binary-coded traits) were excluded, and multiple testing was adjusted using Bonferroni correction.

### Aortic valve area polygenic score calculations

Summary statistics from the aortic valve area GWAS were used to compute a polygenic score on the remaining set of unrelated European-ancestry subjects in the UK Biobank (n=732 aorthopathy cases and 310,546 subjects without a history of aorthopathy). Initially, SNPs with ambiguous alleles (AT or CG), or in linkage disequilibrium with the local SNP with the smallest *p*-value were removed (the SNP with the smallest *p*-value within a 250□kb window is retained, and all neighbors with a linkage disequilibrium *r*^2^□>□0.1 are removed; a step known as clumping). Also, SNPs within the major histocompatibility complex region (chromosome 6, 26–33□Mb) were omitted, and only markers with *p*<0.05 in the aortic valve size GWAS were included in the final polygenic score.

### Genetic correlation and functional mapping

A SNP-based heritability estimate was obtained by LD Score Regression ^23^, submitting the aortic valve GWAS results to LDHub ^24^. In addition, the LDHub platform provided genetic correlation values between the aortic valve summary statistics and a total of 855 human traits from previously released data (597 phenotypes analyzed by the Neale lab on unrelated European-ancestry subjects in the UK Biobank (http://www.nealelab.is/uk-biobank) and 258 phenotypes available mostly from published academic literature). Functional mapping and annotation of genetic associations was conducted through the FUMA platform ^17^.

## Data Availability

UK Biobank clinical & imaging data is available to any qualified researcher at https://www.ukbiobank.ac.uk/

## ACKNOWLEDGEMENTS

We thank Seung-Pyo Lee for his initial contributions to the start of this project, and Dr. Henry Chubb and Dr. Shiraz Maskatia for their contributions in annotating MRI data. We thank the Brin-Wojcicki foundation for support (EA, JRP). JRP was funded by the NIH (K99HL130523). BK is supported by a British Heart Foundation personal chair. This work was supported in part by the Mobilize Center, a National Institutes of Health Big Data to Knowledge (BD2K) Center of Excellence supported through Grant U54EB020405 (J.F., M.F.), the National Science Foundation (NSF) Graduate Research Fellowship under No. DGE-114747 (P.V.), Joseph W. and Hon Mai Goodman Stanford Graduate Fellowship (P.V.). We gratefully acknowledge the support of DARPA under No. FA87501720095 (D3M), ONR under No. N000141712266 and No. N000141410102, and members of the Stanford DAWN project: Intel, Microsoft, Teradata, Google, Facebook, and VMware. The U.S. Government is authorized to reproduce and distribute reprints for Governmental purposes notwithstanding any copyright notation thereon. Any opinions, findings, and conclusions or recommendations expressed in this material are those of the authors and do not necessarily reflect the views, policies, or endorsements, either expressed or implied, of DARPA, NIH, ONR, or the U.S. Government.

## AUTHOR CONTRIBUTIONS

J.P. conceived the initial study and annotated validation data. J.A.F., P.V., V.S.C., K.X. and H.T wrote code and conducted experimental analysis of machine learning models. H.T. and K.X. handled data preprocessing. A.C.-P., J.A.F., P.V., M.F., E.A., C.R., and J.P. contributed ideas and experimental designs. A.C.-P. contributed to statistical analysis and prepared the first manuscript draft. All authors contributed to writing and approved the final version of the manuscript.

